# Neural asymmetry during memory encoding and its association with markers of preclinical Alzheimer’s Disease

**DOI:** 10.1101/2021.06.02.21258020

**Authors:** Jinghang Li, Elizabeth J. Mountz, Akiko Mizuno, Ashti M Shah, Andrea Weinstein, Ann D. Cohen, William E. Klunk, Beth E. Snitz, Howard J. Aizenstein, Helmet T. Karim

**Author notes:** **Corresponding author**: Helmet T. Karim, PhD, Assistant Professor of Psychiatry and Bioengineering, University of Pittsburgh, Pittsburgh PA, 15213. These authors contributed equally to this work (Jinghang Li, Elizabeth J. Mountz).

## Abstract

**Background:** Alzheimer’s Disease (AD) is the most common form of dementia and is characterized by cognitive dysfunction that impacts daily functioning. Beta-amyloid (Aβ) is a cytotoxic protein that deposits in the brain many years prior to the onset of cognitive dysfunction. The preclinical period is a stage of AD in which significant pathology is present without clinical symptoms. Aβ has been shown to deposit asymmetrically early in the AD trajectory, which has shown to have functional consequences (e.g., asymmetric hypometabolism). We aimed to investigate whether markers of AD and cognitive function were correlated with neural activation asymmetry during memory encoding tasks.

**Methods:** We recruited participants who were cognitively normal to undergo functional magnetic resonance imaging (fMRI), positron emission tomography (PET) imaging, and cognitive testing. We conducted analyses to identify regions of significant activation during a well-established face-name pair memory encoding task, and to identify regions of significant asymmetry. We then computed hemispheric asymmetry (negative/positive values indicate left/right asymmetry, respectively) and absolute asymmetry (greater values indicate greater asymmetry in either hemisphere) and investigated their associations with age, sex, education, global cerebral amyloid, global cerebral metabolism, memory encoding task performance, white matter hyperintensities, and multiple domains of cognitive function.

**Results:** We identified expected regions of significant activation, including the hippocampus, and identified four regions with significant left-hemisphere asymmetry: superior medial frontal gyrus, middle frontal gyrus, supplemental motor area, and medial orbitofrontal gyrus, and two regions with significant right hemisphere asymmetry: putamen and ventral posterolateral nucleus of the thalamus. We found that greater left-hemisphere asymmetry in the middle frontal gyrus was correlated with greater global cerebral glucose metabolism. We also found that better performance in memory, learning, and executive attention was associated with greater absolute symmetry in the thalamus, while better visuospatial performance was associated with greater putamen absolute symmetry.

**Discussion:** Functional asymmetry is correlated with functional markers (e.g., glucose metabolism) in older cognitively normal adults and may reflect metabolic and cognitive changes. Longitudinal studies may help us better understand these associations and the causal impact of neural activation asymmetry.

## INTRODUCTION

Alzheimer’s Disease (AD) is the leading cause of dementia, and is characterized by loss of memory and other cognitive functions earlier in the clinical disease process [1]. There is no known cure for AD, so understanding the mechanism of progression is key to early intervention. AD progression is associated with a gradual accumulation of neurofibrillary tangles, white matter hyperintensities (WMHs) [2], and amyloid beta (Aβ) deposition [3]. Since the development of radioactive tracers like Pittsburgh Compound B (PiB), we can measure onset of AD pathology prior to onset of clinical symptoms [1]. This period in which a person has significant Aβ but no cognitive impairment is known as the preclinical stage of AD [1].

Previous research shows that Aβ may accumulate asymmetrically and is associated with asymmetric hypometabolism [4]. Other effects of significant Aβ burden include reduced global glucose metabolism [5], atrophy [6], and functional changes in neuronal activity [4]. Generally, greater disease pathology in aging has also been shown to be associated with compensatory increases in neural activation and increased spread of activation. This phenomenon is referred to as dedifferentiation, defined as a loss of brain functional specificity [7]. Dedifferentiation is a postulated mechanism whereby additional neural resources are needed with accumulating cytotoxic protein and brain atrophy, which preserves behavioral function (e.g., cognitive function) for longer periods [7, 8].

Left-right hemispheric asymmetry is one measure of dedifferentiation; however, the brain is naturally asymmetric such as left-hemispheric language dominance [9, 10]. Cross-sectional studies observing associations between extent of pathology and extent of functional asymmetry may help us better understand pathology-induced changes in neural activation [11, 12]. However, many studies that have delineated the age or pathology related dedifferentiation in cross-sectional settings by comparing older to younger adults. For example, one study found that greater symmetry of activation in the prefrontal regions was associated with older age in episodic memory encoding tasks, while only left lateralized activation was observed in younger adults performing the same tasks [13]. In AD, extent of cerebral asymmetry is associated with lower cognitive function and greater disease severity [11]. Left hemispheric atrophy in cerebral volume specifically has been consistently identified by multiple studies in AD, and has been associated with worse prognosis, including greater AD pathology and lower cognitive function [11, 12]. These may indicate that compensatory mechanisms have an upper limit, and asymmetry is not necessarily inherently a marker of disease.

To our knowledge, previous studies have not evaluated neural activation asymmetry in the preclinical phases of AD. We measured neural activity during a well-established memory encoding task [14] and identified regions of significant asymmetry. We then evaluated associations between AD factors and asymmetric activations. We hypothesized that greater left-lateralized activation would be associated with greater AD-related pathology (less Aβ deposition and WMHs and greater global glucose metabolism) and task performance.

## METHODS

### Participants and Study Design

We recruited 87 participants who were cognitively normal to undergo magnetic resonance (MR) imaging and positron emission tomography (PET) imaging. We included participants who were >65 years of age, fluent in English, and whose cognitive function was normal. Clinical impairment (MCI) was assessed using methods consistent with those used at the University of Pittsburgh Alzheimer Disease Research Center. The criteria for MCI included performance on the neurocognitive assessments below expectations (>1 standard deviation below age and education adjusted norms) on at least 2 tests within the same domain, or at least three tests across domains, reports from the participant of changes or concerns about memory or cognition, and behavioral observations by staff. Results were reviewed by blinded neuropsychologists (BES) and geriatric psychiatrists (WEK and HJA), and each clinical diagnosis was reached by consensus. We excluded participants with diagnoses of mild cognitive impairment or dementia, history of major psychiatric or neurologic disorders, unstable medical conditions, sensory deficits that would preclude cognitive testing, contraindications to MRI, or who were on medications that could affect cognitive function. This study was approved by the University of Pittsburgh Institutional Review Board, and participants gave written informed consent prior to participating in the study. In this analysis, we focus on baseline visits (as this was part of a larger longitudinal study).

### Neurocognitive Assessments

All participants went through a series of neuropsychological tests used by the University of Pittsburgh Alzheimer Disease Research Center (ADRC) to assess cognitive function. For each neuropsychological test conducted, we combined the test scores into domain composite z-scores calculated using the sample averages, which reflected cognitive function in memory (Logical Memory 1 & 2 from the Wechsler Memory Scale - Revised, Modified Rey-Osterreith [R-O] Figure recall, Consortium to Establish a Registry for Alzheimer Disease [CERAD] Word List Memory Test); visuospatial function (Wechsler Adult Intelligence Scale-Revised [WAIS-R] Block Design, Modified R-O Figure copy); language (Animal and Letter Fluency, the 60-Item Boston Naming Test); and executive attention (Trail Making Test A and B, Clock Drawing, WAIS-R Maximum Digit Span Forward and Backward, paper and pencil version of the Stroop Interference Score, and WAIS-R Digit Symbol Substitution) [15][16]. Additionally, the memory domain was split into learning (immediate recall of the listed tests) and retrieval (delayed recall) to isolate the role of the prefrontal cortex in delayed memory retrieval. The z-scores of all tests within each domain were averaged to calculate a standard cognitive domain score.

### MRI Acquisition

MR data was collected on a 3T Siemens Trio scanner using a 12-channel head coil located at the MR Research Center at the University of Pittsburgh. We collected a high resolution structural T1-weighted magnetization prepared rapid gradient echo (MPRAGE) with repetition time (TR)=2300ms, echo time (TE) = 900 ms, flip angle = 9°, FOV = 256 × 224 mm, 176 slices, and 1-mm isotropic voxels. We acquired a T2*-weighted blood oxygen-level-dependent (BOLD) during the face-name pair encoding task with gradient-echo echo-planar imaging with TR = 2000 ms, TE = 34 ms, FOV = 128 × 128, 28 slices, and 2 × 2 × 4 mm voxel size. T2-weighted fluid-attenuated inversion recovery (FLAIR) was acquired with TR = 9160 ms, TE = 90 ms [effective], inversion time (TI) = 2500 ms, FOV = 212 × 256, 48 slices, and 1 × 1 × 3 mm resolution with no slice gap. MR scanning lasted approximately one hour as other sequences were run (not reported).

### Face-Name Pair Memory Encoding Task

The face-name memory encoding task is known to reliably activate areas like the hippocampus and dorsolateral prefrontal cortex [17]. The face-name task has been widely studied in AD progression as a memory encoding task [14]. For the face-name task, participants are first shown two face-name pairs outside of the MR scanner. These serve as the control face-name pairs. During the task in the scanner, using a block design (8 face-name pairs for each block), participants were shown either the control face-name pairs or novel face-name pairs and were asked to make subjective judgements about whether the name shown “fits” the face (Figure 1). While in the scanner, new face-name pairs were shown to the participants and were actively encoded within the scanner. Blocks of familiar and new face-name pairs were alternated. The new face-name pairs that the participants encountered in the scanner were the novel state, and the familiar faces-name pairs were the control state. Both novel and control state activation values were measured with reference to a resting state, during which the participant focused on white crosshairs on a dark screen. Outside of the scanner, participants were shown those same faces and two potential names and were asked to identify the one they saw in the scanner. Post-scan accuracy was computed as the percent of face-name pairs accurately encoded during the face-name task.

**Figure 1.**
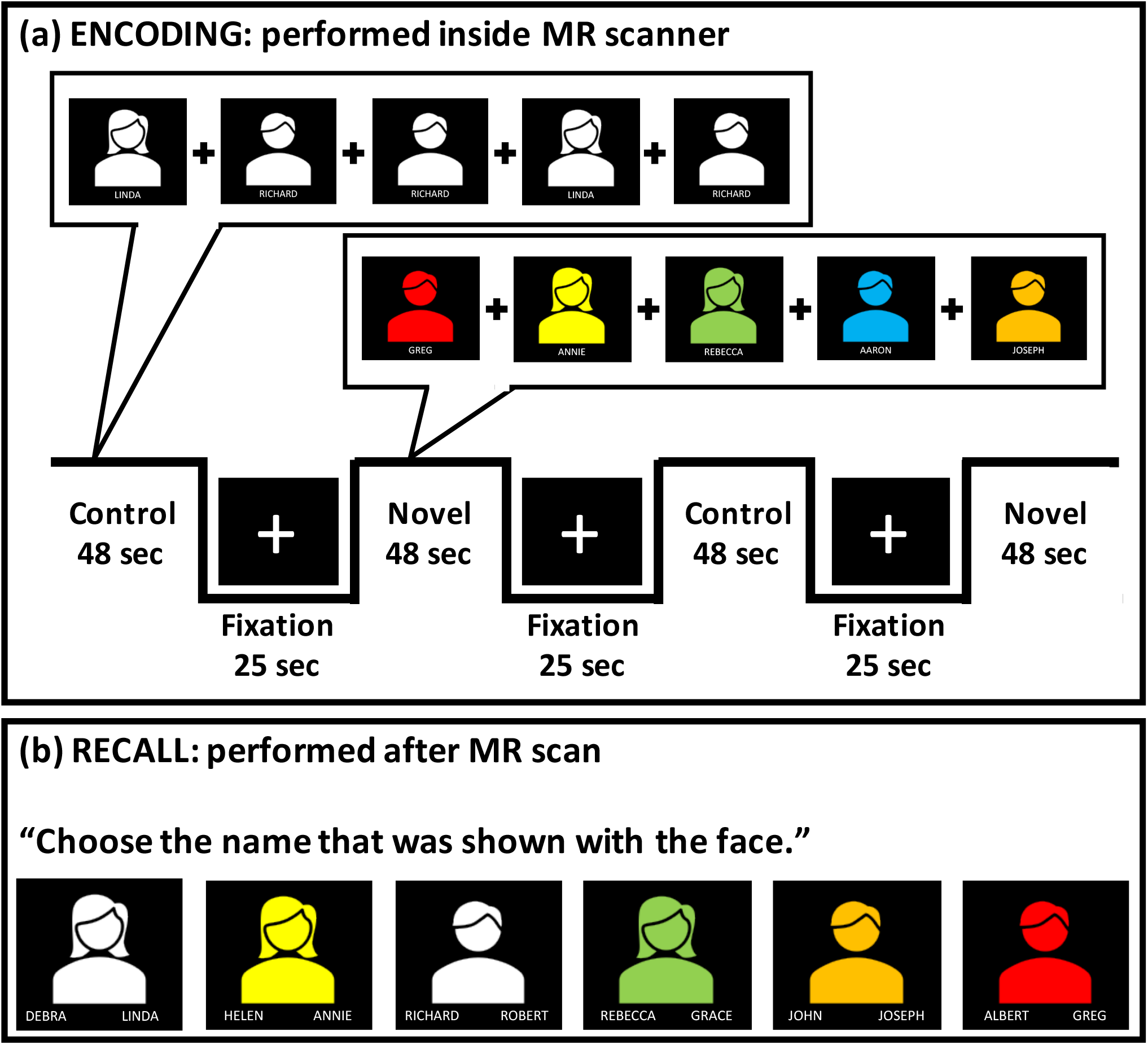
The face-name pair memory encoding task. Prior to the task, participants were shown two control face-name pairs (one male and one female, shown in white), which they were asked to remember. During the fMRI scan, participants were shown both novel (shown in color) and familiar (i.e., control shown in white) face-name pairs, so that novel pairs were actively encoded in the scanner (a). They were instructed to remember the face-name pairs and to answer whether the name fits the face being shown (they were also told that they were going to be tested after the scan). Activation values during both novel and control conditions were measured relative to activation values during the fixation periods, during which the participant focuses on white crosshairs. After the scan, participants were shown the faces seen in the scanner (total of 50 faces) and asked to choose between two names (one of which is the correct name), assessing encoding performance (b) [4]. Approximately 21 cues were non-white faces (42%) and half were female sex.

### Structural Image Processing

Structural images were co-registered to the MPRAGE then segmented using SPM12 multispectral segmentation that generates a deformation field which can be used to normalize functional imaging data to MNI space. We then threshold probability maps of gray matter, white matter, and cerebrospinal fluid by 0.1 and conducted image filling and closing using MATLAB functions to generate an intracranial volume (ICV) mask. MPRAGE images were skull stripped to improve co-registration with functional imaging data. Participants’ WMHs were quantified using a semi-automatic fuzzy connectedness algorithm that segments T2-FLAIR images [18]. All segmentations were reviewed manually to ensure appropriate segmentation.

### Functional Image Processing

The functional image processing was done using SPM12. We conducted motion correction, co-registration to the skull stripped MPRAGE, normalization to MNI space (using deformation field generated in structural processing), and then spatial smoothing using a Gaussian kernel of full width at half-maximum of 8mm. The novel and control condition effects were modeled using boxcars of onsets and durations convolved with the hemodynamic response function using a general linear model. We included a high-pass filter (1/128Hz) to account for drift and an autoregressive [AR(1)] model to account for serial correlations due to aliased biorhythms or unmodeled activity. The contrast novel minus control was generated for group level analyses.

### PET Data Acquisition and Analysis

PiB-PET acquisition and analysis utilized previously described and validated approach [19]. PiB was injected intravenously (12-15mCi, over 20s, specific activity 1-2Ci/μmol) and PET acquisition occurred over the 50-70 min post-injection period. MR images were used for co-registration and region of interest definitions which utilized FreeSurfer segmentation. Standardized uptake value ratios (SUVR) were calculated as the ratio of regional PiB to that of cerebellar gray matter correcting for cerebral atrophy. Global PiB was calculated as the average of nine regions defined within FreeSurfer: anterior cingulate, anterior ventral striatum, superior frontal, orbitofrontal, insula, lateral temporal, parietal, posterior cingulate, and precuneus. We divided these participants into groups of high and low PiB using a median split for statistical power.

Global glucose metabolism was measured using fluorodeoxyglucose (FDG) tracer and PET imaging, which we have described in past work [5]. A similar approach was used to compute global cerebral glucose metabolism as we did for PiB. FDG SUVR was calculated for each of the nine regions at 40-60 minutes post-injection and corrected for cortical atrophy. Global cerebral metabolism was the average of the nine regions.

### Statistical Analysis

We used Statistical non-Parametric Mapping (SnPM) to conduct group level analyses, which computes non-parametric p-values using permutation testing (10,000 permutations used). We first conducted a paired t-test between novel and control conditions in all participants to identify regions of significant activation during the novel compared to the control condition. We used a cluster forming threshold of *p*=0.001 and corrected for multiple comparisons by controlling the cluster family-wise error (FWE) rate at 0.05. This analysis excluded 17 participants whose coverage was poor in the motor cortex, supplemental motor area, or frontal cortex. Our analysis included 70 participants with full coverage, but data was excluded pairwise for all analyses.

We then generated a mask of voxels that were significantly active in at least one hemisphere during the face-name task. We divided all functional clusters using structural definitions from the automated anatomic labeling (AAL3) atlas [20]. We extracted activation (novel-control condition) in each region on the left and right hemisphere (note that all regions have similar number of voxels in the left and right hemisphere). To identify regions that have significant asymmetry, we conducted paired t-tests on the novel-control values of each region between the left and right hemisphere. We controlled the false-discovery rate (FDR) at 0.05 [21].

For regions with significant asymmetry, we computed two markers of asymmetry [22]. We first computed directional asymmetry, which is the difference of the left and the right average activation divided by the sum of the absolute value of the left and the right average activation (novel-control). Positive values indicate greater left-hemisphere dominance while negative values indicate greater right-hemisphere dominance and zero indicates perfect symmetry. We then computed the absolute value of asymmetry such that positive values reflect asymmetry in either the left or right hemisphere and zero indicates perfect symmetry. Absolute asymmetry reflects asymmetry in either hemisphere and represents symmetry vs. lack of symmetry.

In regions with significant asymmetry, we tested their association with factors related to AD. To assess the relationship between AD factors and functional asymmetry in each region, we tested the Pearson’s correlation between asymmetry or absolute asymmetry and the following: task performance (as indicated by post-scan retrieval), cerebral glucose metabolism, white matter hyperintensities (WMH), age, and education. Additionally, we adjusted the WMH correlation test for each participant’s total intracranial volume. We conducted independent t-tests (two-tailed) on asymmetry or absolute asymmetry between PiB groups and sex. Within each analysis, we controlled the False Discovery Rate at 0.05 across all regions. All analyses included only available data (i.e., removed incomplete data with no imputation). We repeated these analyses across domains of cognitive function correcting for multiple corrections by controlling the FDR at 0.05.

## RESULTS

Table 1 shows group differences between high and low PiB (median split). We found that the high PiB group had fewer years in education compared to the low PiB group but did not differ in other values including cognitive function.

**Table 1.**
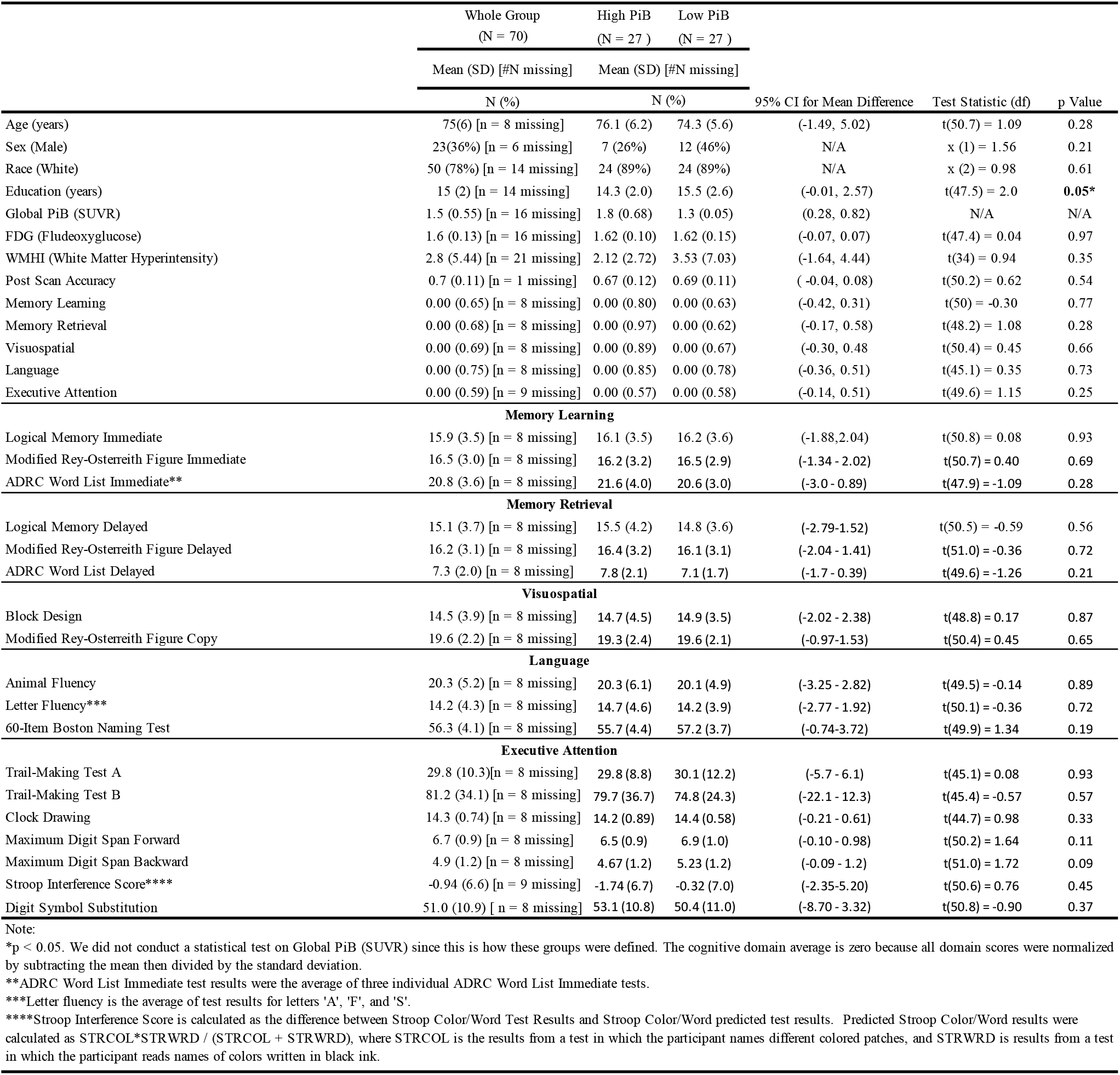
Demographics and AD risk factors measurement*.

We first identified regions of statistically significant activation during the face-name task using a voxel-wise whole brain paired t-test (see Figure 2a and Table 2). We identified significant activation in the pregenual anterior cingulate, middle and posterior cingulate, inferior frontal gyrus (inferior operculum, orbital, triangular, and medial orbital), middle frontal gyrus, superior frontal gyrus, superior medial frontal gyrus, orbitofrontal gyrus (anterior, lateral, and posterior), amygdala, hippocampus, insula, parahippocampus, putamen, red nucleus, substantia nigra (pars reticulata), nuclei of the thalamus (magnocellular portion of the mediodorsal nucleus), medial pulvinar nucleus, ventral lateral nucleus, and ventral posterolateral nucleus), olfactory/rectus gyrus, temporal gyrus (inferior and superior pole), fusiform gyrus, visual cortex (calcarine, cuneus, lingual inferior occipital, and precuneus gyrus), supplemental motor area, cerebellar lobe IV-V (sensorimotor), and Cerebellar lobe VI (mental imagery).

**Figure 2.**
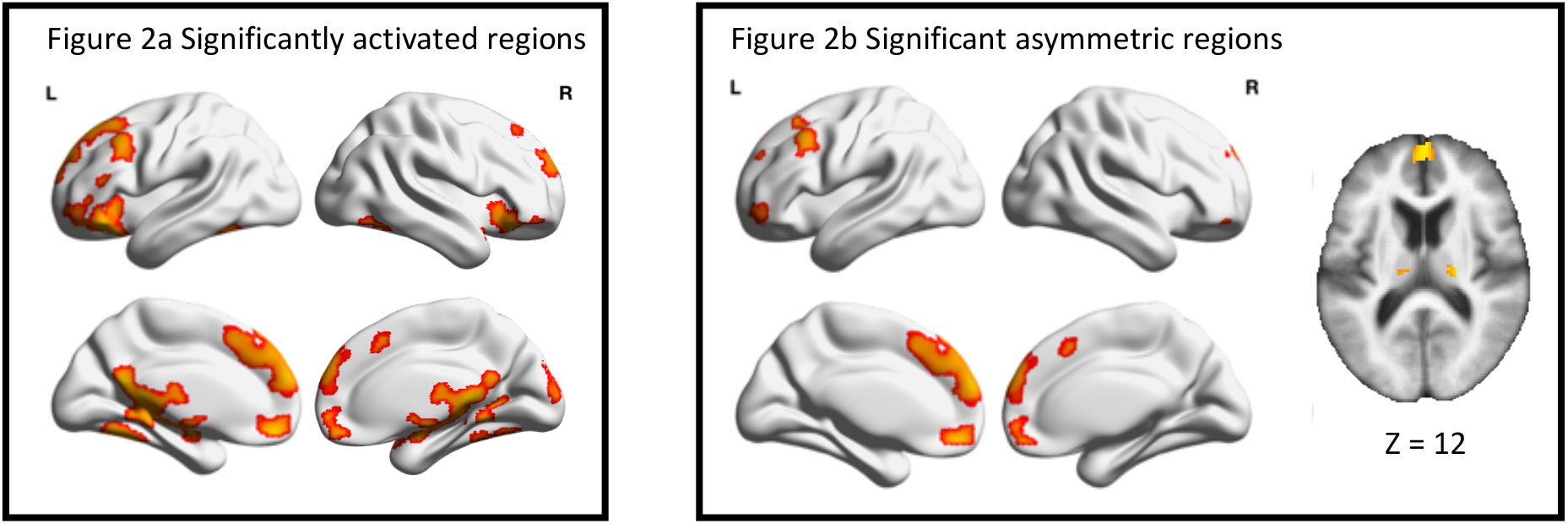
a) The areas that were significantly activated during the face-name memory encoding task (result of the voxel-wise paired t-test). b) The areas that were significantly asymmetric during the face-name memory encoding task. Generated using BrainNet viewer on a smoothed average brain template. In the last column, xjview was used to show thalamus and putamen regions.

**Table 2.** Regions significantly activated by the memory encoding task (novel>control). Separate clusters are separated by a gray bar. Each region was separated using the automated anatomical labeling 3 (AAL3) atlas. All clusters were significant after controlling the voxel-wise family-wise error (FWE) < 0.05. The max t-statistic (paired t-test), cluster size (in voxels), and MNI coordinates (x, y, z) are reported.

| Side | Region | Num. Voxels | T-statistic (max) | x MNI | y MNI | z MNI |
| --- | --- | --- | --- | --- | --- | --- |
| Left | Fusiform | 423 | 5.8 | -36 | -52 | -18 |
| Left | Inferior Frontal (orbital) | 394 | 5.8 | -44 | 30 | -12 |
| Right |  | 278 | 5 | 42 | 26 | -12 |
| Left | Precuneus | 311 | 5.9 | -4 | -54 | 20 |
| Right |  | 115 | 4.9 | 6 | -44 | 8 |
| Left | Hippocampus | 255 | 5.5 | -22 | -34 | -4 |
| Right |  | 74 | 5 | 20 | -10 | -12 |
| Right | Lingual | 253 | 5.2 | 4 | -38 | 2 |
| Left |  | 187 | 5.7 | -12 | -40 | -2 |
| Right | Insula | 239 | 4.3 | 48 | 16 | -2 |
| Left |  | 171 | 4.4 | -26 | 18 | -20 |
| Bilateral | Vermis_4_5 | 230 | 5.8 | 0 | -42 | 2 |
| Left | Orbitofrontal (posterior) | 220 | 6.8 | -42 | 30 | -16 |
| Right |  | 168 | 5.9 | 42 | 28 | -14 |
| Left | Cerebellum IV/V | 210 | 5.8 | -6 | -40 | 0 |
| Right |  | 72 | 4 | 10 | -52 | -6 |
| Left | Cerebellum VI | 126 | 5 | -28 | -56 | -18 |
| Bilateral | Vermis VI | 120 | 4.7 | -2 | -74 | -12 |
| Left | Posterior Cingulate | 113 | 5.9 | -4 | -42 | 8 |
| Right |  | 72 | 5.9 | 2 | -40 | 8 |
| Right | Ventral Posterolateral Nucleus Thalamus | 107 | 4.9 | 20 | -18 | 6 |
| Left | Calcarine | 98 | 5.4 | -6 | -44 | 6 |
| Right |  | 77 | 3.9 | 28 | -52 | 4 |
| Left | Orbitofrontal (lateral) | 95 | 6.6 | -44 | 30 | -14 |
| Right |  | 20 | 4.8 | 42 | 36 | -16 |
| Right | Ventral Lateral Thalamus | 91 | 4.4 | 20 | -16 | 4 |
| Left | Parahippocampus | 82 | 5.1 | -22 | -36 | -6 |
| Left | Orbitofrontal (anterior) | 77 | 6.3 | -36 | 36 | -16 |
| Right |  | 34 | 5.2 | 40 | 34 | -14 |
| Right | Medial Pulvinar Nucleus Thalamus | 59 | 4.2 | 14 | -30 | 2 |
| Left |  | 40 | 3.8 | -6 | -22 | 12 |
| Right | Amygdala | 57 | 4.9 | 24 | -6 | -16 |
| Left |  | 36 | 5.4 | -22 | -8 | -16 |
| Right | Superior Temporal Pole | 53 | 4.5 | 44 | 18 | -18 |
| Left |  | 25 | 4.6 | -48 | 22 | -12 |
| Right | Inferior Frontal (triangular) | 50 | 4.1 | 48 | 20 | 0 |
| Right | Inferior Frontal (operculum) | 47 | 4.4 | 50 | 16 | -2 |
| Left | Inferior Temporal | 40 | 4.8 | -40 | -48 | -18 |
| Left | Medial Mediodorsal Thalamus | 37 | 3.9 | -2 | -18 | 8 |
| Right | Red Nucleus | 29 | 4.2 | 4 | -16 | -12 |
| Right | Putamen | 23 | 3.7 | 28 | -12 | 6 |
| Bilateral | Vermis III | 22 | 4.7 | 0 | -42 | -2 |
| Right | Substantia Nigra (pars reticulata) | 22 | 4.1 | 14 | -10 | -10 |
| Right | Fusiform | 193 | 5.9 | 36 | -46 | -18 |
| Right | Inferior Temporal | 46 | 4.3 | 42 | -60 | -12 |
| Right | Inferior Occipital | 35 | 5.3 | 38 | -64 | -12 |

| Table 2 (Continued) |  |  |  |  |  |  |
| --- | --- | --- | --- | --- | --- | --- |
| Side | Region | Num. Voxels | T-statistic (max) | x MNI | y MNI | z MNI |
| Left | Medial Frontal (orbital) | 269 | 5.7 | -4 | 46 | -14 |
| Right |  | 131 | 4.9 | 2 | 48 | -14 |
| Left | Rectus gyrus | 32 | 5.8 | -2 | 46 | -16 |
| Right |  | 29 | 5.4 | 2 | 50 | -16 |
| Left | Cuneus | 151 | 5.4 | 0 | -88 | 36 |
| Right |  | 42 | 4.5 | 6 | -94 | 20 |
| Left | Superior Medial Frontal | 1224 | 6.5 | -2 | 60 | 22 |
| Right |  | 380 | 5.3 | 2 | 62 | 16 |
| Left | Superior Frontal | 527 | 5.1 | -22 | 30 | 54 |
| Right |  | 299 | 4.4 | 14 | 56 | 28 |
| Left | Supplemental Motor | 173 | 4.4 | -4 | 22 | 46 |
| Right |  | 29 | 3.7 | 2 | 18 | 52 |
| Left | Anterior Cingulate (pregenual) | 59 | 4.3 | -2 | 50 | 14 |
| Right | Middle Frontal | 27 | 3.6 | 26 | 58 | 26 |
| Left | Middle Frontal | 435 | 6.2 | -28 | 24 | 54 |
| Left | Inferior Frontal (triangular) | 247 | 4.8 | -52 | 24 | 30 |
| Left | Inferior Frontal (operculum) | 48 | 4.9 | -52 | 22 | 32 |

We then found that there were six regions (after controlling FDR at 0.05) that had significant asymmetry (Table 3). Several regions had significant left hemisphere asymmetry: superior medial frontal gyrus, middle frontal gyrus, supplemental motor area, and medial orbitofrontal gyrus, while other regions had significant right hemisphere asymmetry: putamen and ventral posterolateral nucleus of the thalamus (see Figure 2b).

**Table 3.** All the regions below were significantly activated during DSST. Additionally, regions that were significantly and asymmetrically activated are bolded. The t-statistics, p-value and FDR corrected q-value are reported.

| Region | t-test, p-value, FDR corrected q-value<br>(Positive=Left>Right) |
| --- | --- |
| Superior Medial Frontal | **t(69) = 6.1, p = 0, q = 0** |
| Medial Frontal (orbital) | **t(69) = 4.1, p = 0, q = 0.002** |
| Middle Frontal | **t(69) = 3.1, p = 0.002, q = 0.029** |
| Putamen | **t(69) = -3.0, p = 0.004, q = 0.032** |
| Supplemental Motor | **t(69) = 2.9, p = 0.005, q = 0.036** |
| Ventral Posterolateral Nucleus Thalamus | **t(69) = -2.8, p = 0.007, q = 0.04** |
| Cerebellum IV/V | t(69) = 2.4, p = 0.018, q = 0.083 |
| Rectus gyrus | t(69) = 2.4, p = 0.019, q = 0.083 |
| Precuneus | t(69) = 2.3, p = 0.026, q = 0.088 |
| Inferior Occipital | t(69) = -2.3, p = 0.026, q = 0.088 |
| Cerebellum VI | t(69) = 2.2, p = 0.033, q = 0.089 |
| Middle Cingulate | t(69) = 2.2, p = 0.028, q = 0.088 |
| Superior Frontal | t(69) = 2.2, p = 0.03, q = 0.088 |
| Orbitofrontal (anterior) | t(69) = 2.1, p = 0.038, q = 0.089 |
| Orbitofrontal (lateral) | t(69) = 2.1, p = 0.038, q = 0.089 |
| Superior Temporal Pole | t(69) = -2.0, p = 0.047, q = 0.103 |
| Inferior Temporal | t(69) = -1.8, p = 0.077, q = 0.154 |
| Ventral Lateral Thalamus | t(69) = -1.8, p = 0.079, q = 0.154 |
| Inferior Frontal (operculum) | t(69) = 1.7, p = 0.093, q = 0.171 |
| Inferior Frontal (triangular) | t(69) = 1.6, p = 0.121, q = 0.202 |
| Cuneus | t(69) = -1.6, p = 0.108, q = 0.189 |
| Anterior Cingulate (pregenual) | t(69) = 1.5, p = 0.135, q = 0.215 |
| Medial Mediodorsal Thalamus | t(69) = 1.4, p = 0.181, q = 0.264 |
| Medial Pulvinar Nucleus Thalamus | t(69) = -1.4, p = 0.155, q = 0.237 |
| Parahippocampus | t(69) = 1.2, p = 0.24, q = 0.336 |
| Red Nucleus | t(69) = -0.9, p = 0.348, q = 0.468 |
| Insula | t(69) = -0.8, p = 0.419, q = 0.543 |
| Inferior Frontal (orbital) | t(69) = 0.7, p = 0.508, q = 0.613 |
| Orbitofrontal (posterior) | t(69) = 0.7, p = 0.482, q = 0.603 |
| Hippocampus | t(69) = 0.4, p = 0.679, q = 0.792 |
| Posterior Cingulate | t(69) = -0.3, p = 0.772, q = 0.872 |
| Calcarine | t(69) = 0.2, p = 0.866, q = 0.918 |
| Amygdala | t(69) = -0.2, p = 0.806, q = 0.881 |
| Lingual | t(69) = 0.1, p = 0.939, q = 0.958 |
| Fusiform | t(69) = -0.1, p = 0.958, q = 0.958 |

We conducted statistical analyses between asymmetry and multiple factors related to AD including global cerebral metabolism, global amyloid, WMH, task performance, age, sex, and education (Table 4). We found that greater global cerebral metabolism was correlated with greater left-hemisphere asymmetry in the middle frontal gyrus (Figure 3). We found no other significant associations. We found no association between absolute asymmetry and factors related to AD, including global cerebral metabolism, global amyloid, WMH, task performance, age, sex, and education (Table 5).

**Table 4.** Associations between functional Asymmetry and AD Factors. We report the statistical test (1-Pearson’s correlation, 2-Independent t-test, and 3-Partial Pearson’s Correlation), degrees of freedom, and p-values as well as the adjusted q-value from the FDR correction (correcting across 6 regions per set of 6 tests).

| Region | Global Cerebral Metabolism<sup>1</sup><br>(FDG PET SUVR) | Global Amyloid<sup>2</sup><br>(PiB SUVR) | WMHI<sup>3</sup><br>(normalized by ICV) | Memory Encoding Task Performance<sup>1</sup><br>(Accuracy) | Age<sup>1</sup><br>(years) | Sex<sup>2</sup><br>(reference group Female) | Education<sup>1</sup><br>(years) |
| --- | --- | --- | --- | --- | --- | --- | --- |
| Putamen | r(52) = 0.287<br>p = 0.035*<br>q = 0.105 | t(48.42) = 0.118<br>p = 0.906<br>q = 0.993 | r(60) = 0.123<br>p = 0.342<br>q = 0.943 | r(67) = 0.175<br>p = 0.149<br>q = 0.298 | r(60) = -0.048<br>p = 0.707<br>q = 0.707 | t(54.04) = 1.909<br>p = 0.061*<br>q = 0.185 | r(54) = -0.105<br>p = 0.442<br>q = 0.531 |
| Thalamus ventral posterolateral nucleus | r(52) = 0.247<br>p = 0.071<br>q = 0.142 | t(49.49) = -0.305<br>p = 0.761<br>q = 0.993 | r(60) = 0.009<br>p = 0.943<br>q = 0.943 | r(67) = 0.253<br>p = 0.036*<br>q = 0.108 | r(60) = -0.240<br>p = 0.060*<br>q = 0.179 | t(40.25) = 1.433<br>p = 0.159<br>q = 0.276 | r(54) = 0.110<br>p = 0.420<br>q = 0.531 |
| Superior medial frontal gyrus | r(52) = 0.128<br>p = 0.355<br>q = 0.355 | t(51.84) = -0.681<br>p = 0.498<br>q = 0.993 | r(60) = -0.245<br>p = 0.055*<br>q = 0.328 | r(67) = 0.273<br>p = 0.023*<br>q = 0.108 | r(60) = -0.307<br>p = 0.015*<br>q = 0.092 | t(33.41) = 1.250<br>p = 0.220<br>q = 0.276 | r(54) = -0.027<br>p = 0.844<br>q = 0.844 |
| Middle frontal gyrus | r(52) = 0.342<br>p = 0.011*<br>**q = 0.068\*** | t(51.86) = -0.137<br>p = 0.890<br>q = 0.993 | r(60) = -0.062<br>p = 0.634<br>q = 0.943 | r(67) = 0.091<br>p = 0.454<br>q = 0.454 | r(60) = -0.154<br>p = 0.231<br>q = 0.347 | t(44.96) = 2.058<br>p = 0.045*<br>q = 0.185 | r(54) = -0.113<br>p = 0.41<br>q = 0.531 |
| Supplemental motor area | r(52) = 0.149<br>p = 0.283<br>q = 0.339 | t(47.70) = 0.439<br>p = 0.662<br>q = 0.993 | r(60) = -0.077<br>p = 0.549<br>q = 0.943 | r(67) = -0.133<br>p = 0.277<br>q = 0.332 | r(60) = 0.116<br>p = 0.368<br>q = 0.441 | t(37.56) = 1.219<br>p = 0.230<br>q = 0.276 | r(54) = -0.151<br>p = 0.267<br>q = 0.531 |
| Medial orbitofrontal gyrus | r(52) = 0.152<br>p = 0.273<br>q = 0.339 | t(49.38) = -0.009<br>p = 0.993<br>q = 0.99 | r(60) = 0.015<br>p = 0.910<br>q = 0.943 | r(67) = 0.151<br>p = 0.214<br>q = 0.321 | r(60) = -0.199<br>p = 0.120<br>q = 0.240 | t(35.19) = -0.360<br>p = 0.721<br>q = 0.721 | r(54) = 0.225<br>p = 0.096<br>q = 0.531 |

**Table 5.** Associations between functional absolute Asymmetry and AD Factors. We report the statistical test (1-Pearson’s correlation, 2-Independent t-test, and 3-Partial Pearson’s Correlation), degrees of freedom, and p-values as well as the adjusted q-value from the FDR correction (correcting across 6 regions per set of 6 tests).

| Region | Global Cerebral Metabolism<sup>1</sup><br>(FDG PET SUVR) | Global Amyloid<sup>2</sup><br>(PiB SUVR) | WMH<sup>3</sup><br>(normalized by ICV) | Memory Encoding Task Performance<sup>1</sup><br>(Accuracy) | Age<sup>1</sup><br>(years) | Sex<sup>2</sup><br>(reference group Female) | Education<sup>1</sup><br>(years) |
| --- | --- | --- | --- | --- | --- | --- | --- |
| Putamen | $r(52) = -0.210$<br>$p = 0.127$<br>$q = 0.381$ | $t(51.88) = 1.71$<br>$p = 0.906$<br>$q = 0.993$ | $r(60) = 0.134$<br>$p = 0.299$<br>$q = 0.477$ | $r(67) = -0.080$<br>$p = 0.515$<br>$q = 0.871$ | $r(60) = 0.147$<br>$p = 0.254$<br>$q = 0.926$ | $t(45.23) = -0.335$<br>$p = 0.739$<br>$q = 0.739$ | $r(54) = -0.026$<br>$p = 0.847$<br>$q = 0.986$ |
| Thalamus ventral posterolateral nucleus | $r(52) = -0.107$<br>$p = 0.440$<br>$q = 0.875$ | $t(51.85) = 1.58$<br>$p = 0.761$<br>$q = 0.993$ | $r(60) = -0.092$<br>$p = 0.477$<br>$q = 0.477$ | $r(67) = -0.179$<br>$p = 0.140$<br>$q = 0.541$ | $r(60) = -0.056$<br>$p = 0.665$<br>$q = 0.926$ | $t(42.67) = -1.538$<br>$p = 0.131$<br>$q = 0.639$ | $r(54) = 0.016$<br>$p = 0.909$<br>$q = 0.986$ |
| Superior medial frontal gyrus | $r(52) = -0.238$<br>$p = 0.083^*$<br>$q = 0.381$ | $t(51.15) = -1.20$<br>$p = 0.499$<br>$q = 0.993$ | $r(60) = 0.098$<br>$p = 0.447$<br>$q = 0.477$ | $r(67) = 0.039$<br>$p = 0.752$<br>$q = 0.903$ | $r(60) = -0.015$<br>$p = 0.905$<br>$q = 0.926$ | $t(39.40) = -0.771$<br>$p = 0.445$<br>$q = 0.739$ | $r(54) = 0.200$<br>$p = 0.139$<br>$q = 0.418$ |
| Middle frontal gyrus | $r(52) = 0.076$<br>$p = 0.584$<br>$q = 0.875$ | $t(50.71) = -0.92$<br>$p = 0.890$<br>$q = 0.993$ | $r(60) = 0.107$<br>$p = 0.410$<br>$q = 0.477$ | $r(67) = 0.068$<br>$p = 0.580$<br>$q = 0.871$ | $r(60) = 0.029$<br>$p = 0.820$<br>$q = 0.926$ | $t(49.53) = 0.450$<br>$p = 0.655$<br>$q = 0.740$ | $r(54) = 0.002$<br>$p = 0.985$<br>$q = 0.986$ |
| Supplemental motor area | $r(52) = 0.002$<br>$p = 0.988$<br>$q = 0.988$ | $t(46.79) = -0.86$<br>$p = 0.663$<br>$q = 0.993$ | $r(60) = -0.142$<br>$p = 0.271$<br>$q = 0.477$ | $r(67) = 0.163$<br>$p = 0.180$<br>$q = 0.541$ | $r(60) = 0.026$<br>$p = 0.841$<br>$q = 0.926$ | $t(36.98) = -0.480$<br>$p = 0.635$<br>$q = 0.739$ | $r(54) = 0.308$<br>$p = 0.021^*$<br>$q = 0.126$ |
| Medial orbitofrontal gyrus | $r(52) = -0.040$<br>$p = 0.771$<br>$q = 0.925$ | $t(51.87) = -1.29$<br>$p = 0.993$<br>$q = 0.993$ | $r(60) = 0.093$<br>$p = 0.470$<br>$q = 0.477$ | $r(67) = 0.009$<br>$p = 0.944$<br>$q = 0.944$ | $r(60) = -0.012$<br>$p = 0.926$<br>$q = 0.926$ | $t(36.22) = -1.268$<br>$p = 0.213$<br>$q = 0.639$ | $r(54) = 0.152$<br>$p = 0.264$<br>$q = 0.527$ |

**Figure 3.**
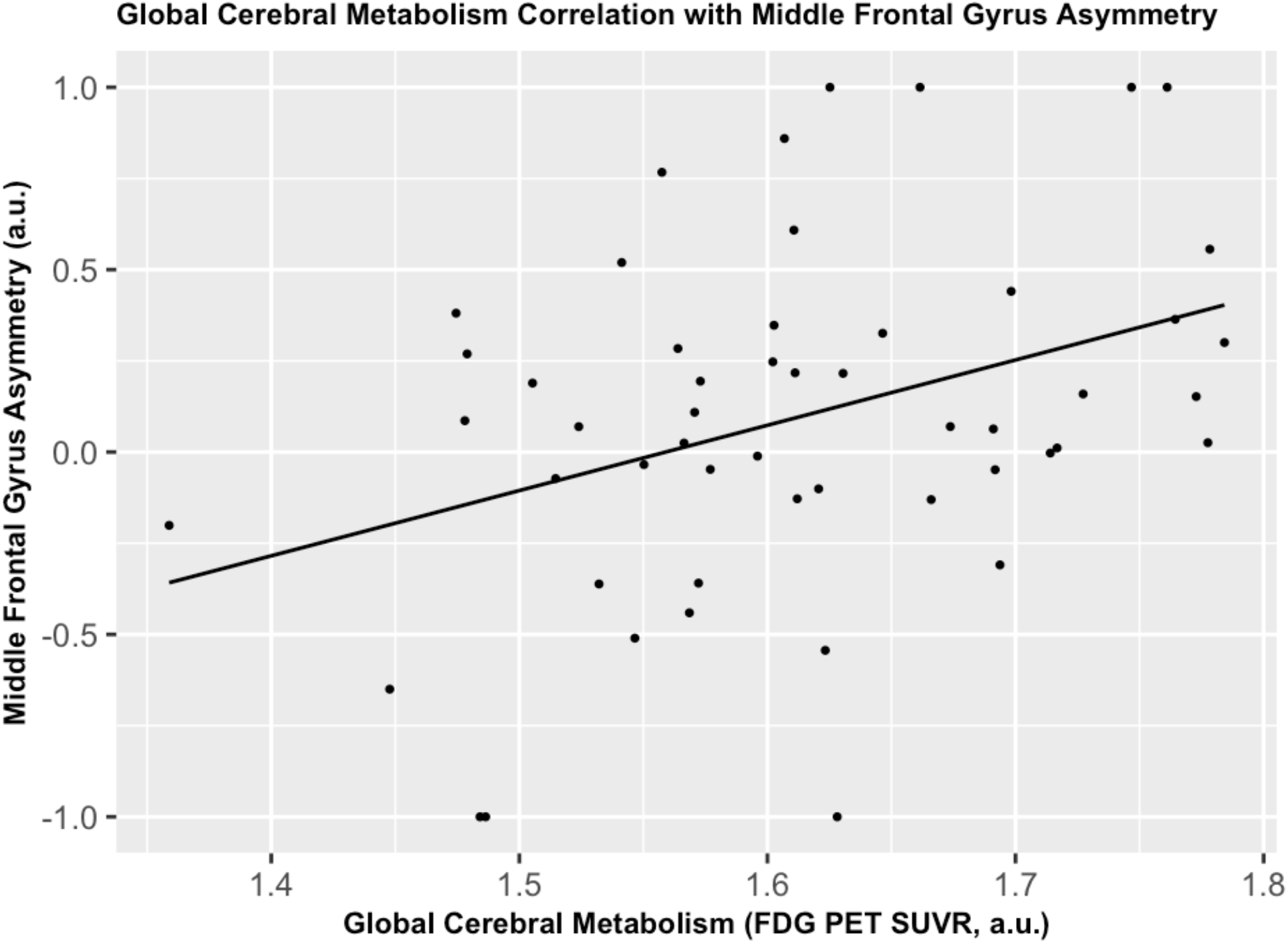
Association between greater global cerebral metabolism (FDG PET SUVR) and greater left-hemisphere asymmetry in the middle frontal gyrus [*r*(52)= 0.342, *p*=0.011, *q*=0.068].

We conducted statistical analyses between asymmetry and multiple cognitive domains (Table 6). We found that asymmetry was not significantly correlated to cognitive function. We repeated our analysis using absolute asymmetry and cognitive domains (Table 7). We found that better cognitive performance in memory learning and executive attention was correlated with lower absolute asymmetry in the ventral posterolateral thalamus (i.e., greater symmetry regardless of the direction [left or right]), while better visuospatial performance correlated with lower absolute asymmetry (i.e., greater symmetry) in the putamen.

**Table 6.** Associations between functional Asymmetry and cognitive domains. We report the Pearson’s correlation coefficients, degrees of freedom, and p-values as well as the adjusted q-value from the FDR correction (correcting across six regions per set of tests). All domains are average z-scores across multiple tests (see methods).

| *Region* | *Memory Learning* | *Memory Retrieval* | *Visuospatial* | *Language* | *Executive Attention* |
| --- | --- | --- | --- | --- | --- |
| *Putamen* | r(60) = -0.119<br>p = 0.358<br>q = 0.755 | r(60) = -0.151<br>p = 0.240<br>q = 0.830 | r(60) = 0.157<br>p = 0.224<br>q = 0.448 | r(60) = -0.245<br>p = 0.055*<br>q = 0.279 | r(59) = 0.009<br>p = 0.943<br>q = 0.943 |
| *Thalamus ventral posterolateral nucleus* | r(60) = 0.023<br>p = 0.861<br>q = 0.915 | r(60) = 0.038<br>p = 0.772<br>q = 0.830 | r(60) = -0.076<br>p = 0.558<br>q = 0.670 | r(60) = -0.007<br>p = 0.957<br>q = 0.957 | r(59) = 0.212<br>p = 0.100<br>q = 0.270 |
| *Superior medial frontal gyrus* | r(60) = 0.161<br>p = 0.212<br>q = 0.755 | r(60) = 0.124<br>p = 0.336<br>q = 0.830 | r(60) = 0.201<br>P = 0.118<br>q = 0.448 | r(60) = 0.215<br>p = 0.093*<br>q = 0.279 | r(59) = 0.109<br>p = 0.401<br>q = 0.602 |
| *Middle frontal gyrus* | r(60) = 0.014<br>p = 0.914<br>q = 0.915 | r(60) = -0.050<br>p = 0.702<br>q = 0.830 | r(60) = 0.050<br>p = 0.697<br>q = 0.697 | r(60) = -0.058<br>p = 0.656<br>q = 0.957 | r(59) = 0.193<br>p = 0.135<br>q = 0.270 |
| *Supplemental motor area* | r(60) = -0.021<br>P = 0.871<br>q = 0.915 | r(60) = -0.058<br>p = 0.656<br>q = 0.830 | r(60) = -0.172<br>p = 0.181<br>q = 0.448 | r(60) = -0.009<br>p = 0.945<br>q = 0.957 | r(59) = 0.045<br>p = 0.729<br>q = 0.875 |
| *Medial orbitofrontal gyrus* | r(60) = 0.114<br>p = 0.378<br>q = 0.755 | r(60) = 0.028<br>p = 0.830<br>q = 0.830 | r(60) = 0.130<br>p = 0.314<br>q = 0.471 | r(60) = 0.117<br>p = 0.364<br>q = 0.728 | r(59) = 0.245<br>p = 0.057*<br>q = 0.270 |

**Table 7.** Associations between functional absolute Asymmetry and cognitive domains. We report the Pearson’s correlation coefficients, degrees of freedom, and p-values as well as the adjusted q-value from the FDR correction (correcting across six regions per set of tests). All domains are average z-scores across multiple tests (see methods).

| *Region* | *Memory Learning* | *Memory Retrieval* | *Visuospatial* | *Language* | *Executive Attention* |
| --- | --- | --- | --- | --- | --- |
| *Putamen* | r(60) = -0.025<br>p = 0.847<br>q = 0.847 | r(60) = 0.002<br>p = 0.989<br>q = 0.997 | r(60) = -0.326<br>p = 0.010*<br>**q = 0.059\*** | r(60) = -0.326<br>p = 0.725<br>q = 0.725 | r(59) = 0.002<br>P = 0.985<br>q = 0.992 |
| *Thalamus ventral posterolateral nucleus* | r(60) = -0.317<br>p = 0.012*<br>**q = 0.072\*** | r(60) = -0.296<br>p = 0.019*<br>q = 0.116 | r(60) = -0.173<br>p = 0.178<br>q = 0.534 | r(60) = -0.173<br>p = 0.279<br>q = 0.558 | r(59) = -0.323<br>p = 0.011*<br>**q = 0.066\*** |
| *Superior medial frontal gyrus* | r(60) = 0.084<br>p = 0.515<br>q = 0.707 | r(60) = 0.034<br>P = 0.792<br>q = 0.997 | r(60) = 0.069<br>p = 0.595<br>q = 0.714 | r(60) = 0.0688<br>p = 0.060*<br>q = 0.279 | r(59) = 0.139<br>p = 0.285<br>q = 0.571 |
| *Middle frontal gyrus* | r(60) = -0.0118<br>p = 0.360<br>q = 0.707 | r(60) = -0.215<br>p = 0.094*<br>q = 0.281 | r(60) = 0.115<br>p = 0.375<br>q = 0.608 | r(60) = 0.115<br>p = 0.09*<br>q = 0.279 | r(59) = -0.001<br>p = 0.992<br>q = 0.993 |
| *Supplemental motor area* | r(60) = -0.070<br>p = 0.589<br>q = 0.707 | r(60) = -0.000<br>p = 0.997<br>q = 0.997 | r(60) = 0.108<br>p = 0.405<br>q = 0.608 | r(60) = 0.108<br>p = 0.476<br>q = 0.580 | r(59) = 0.157<br>P = 0.227<br>q = 0.571 |
| *Medial orbitofrontal gyrus* | r(60) = 0.124<br>P = 0.337<br>q = 0.707 | r(60) = 0.095<br>p = 0.462<br>q = 0.926 | r(60) = 0.033<br>P = 0.800<br>q = 0.800 | r(60) = 0.0328<br>p = 0.483<br>q = 0.580 | r(59) = 0.085<br>p = 0.514<br>q = 0.772 |

## DISCUSSION

We identified regions with significant asymmetric activation during the face-name memory encoding task – four with left-hemispheric asymmetry: superior medial frontal gyrus, middle frontal gyrus, supplemental motor area, and the medial orbitofrontal gyrus and two with right-hemispheric asymmetry: putamen and ventral posterolateral nucleus of the thalamus. We found that greater left hemisphere asymmetry in the middle frontal gyrus was associated with greater cerebral glucose metabolism. We also found that better memory learning and executive attention performance was associated with more symmetric activation in the ventral posterolateral thalamus, while better visuospatial performance correlated with more symmetric activation in the putamen.

Our results are consistent with past work on putamen and thalamus asymmetry. Previous research has demonstrated that the right hemisphere of the putamen is associated with memory and visual imagery processing [23]. Based on this, we can cautiously interpret that more symmetric putamen activation may be associated with a compensatory mechanism (i.e., left hemispheric co-activation) that maintains greater visuospatial performance, whereas normally this activation may be right-hemispheric dominant, which was true in this sample on average. This region may help encode the visual imagery of the faces or even the face-name pairs. One study found that AD patients had greater left hemispheric atrophy compared to the right hemisphere in the thalamus compared to controls [11]. Our study with cognitively normal older adults found that greater thalamus symmetry was associated with greater memory learning and executive attention performance. Since the thalamus was found to have greater right hemisphere asymmetry, we can again speculate that symmetric activations may be associated with compensatory mechanisms that may maintain higher cognitive performance.

We found that greater left dominant activation in the middle frontal gyrus was positively correlated with greater whole-brain cerebral glucose metabolism. Previous research has shown that rightward lateralization of functional activity was associated with greater AD pathology [24, 25]. This region had largely left-hemisphere dominant activation and so its association with a marker of overall metabolism, which is a marker of brain health in the context of AD, suggests that this region may play an important early role in AD. The middle frontal gyrus, which includes the dorsolateral prefrontal cortex, has been largely implicated in cognitive impairment, including AD [26].

By looking at effect sizes and directionality, we observed expected relationships between age, performance, and functional asymmetry. Although there was significant asymmetric activation in all six identified regions during the face-name task, there were no associations between activation asymmetry in these regions and any other AD risk factors. Our findings do not suggest any relationship between Aβ and the functional asymmetry in older cognitively normal individuals. Thus, either Aß is associated with regions that have non-significant asymmetry or may not have a large impact on asymmetry in the preclinical stage of AD.

There is an ongoing question of how disease progression manifests in cortical versus subcortical brain structures [27, 28]. In a longitudinal study, researchers identified significant putamen atrophy and right dominant asymmetry as people age [29]. Additionally, increased ventral posterolateral asymmetry in the thalamus has been noted to be associated with worse cognitive function [11]. These subcortical regions of the brain (i.e., thalamus and putamen) may be impacted earlier in AD [28], when excess Aβ production drives disease progression. One possible explanation could be that significant amyloid burden is interfering with the cortical communication pathway that goes through the subcortical regions like thalamus and putamen. Those with subcortical pathology may therefore have worse outcomes with greater asymmetry, meaning that cognitive reserve has been expended and compensatory mechanisms are no longer able to maintain cognitive function. We found that greater symmetry in the subcortical regions was associated with better cognitive performance. Unlike early-stage AD, late-stage AD is linked closely with cytotoxic protein clearance and may impact cortical structures more than subcortical structures [27]. In late-onset AD, those who have retained leftward dominance despite pathology may have better disease outcomes. Thus, lower glucose metabolism may be correlated with more rightward asymmetry in cortical regions like the middle frontal gyrus.

Our study has several limitations. We analyzed data from the cross-sectional data and did not analyze longitudinal changes in asymmetry, AD factors, and cognitive domain scores during the memory encoding task. Longitudinal studies should address the temporal relationship between these factors by measuring for our predicted model of functional change, in which symmetry increases with age (i.e., more dedifferentiation) and then eventual rightward-dominant asymmetry occurs as Aß and WMHs increase and FDG, task performance, and cognitive domain scores decrease. Furthermore, we analyzed only regions of significant asymmetry during memory encoding – so there may be regions that have varying levels of asymmetry across participants that would not be detected since they on average do not show a significant asymmetry. Future studies should also explore the relationships between AD factors and greater variability in absolute asymmetry by considering all regions of the brain activated during a memory encoding task. To increase our understanding of the associations between asymmetry and AD risk factors, future studies should repeat this study with different functional tasks. We only investigated global differences in Aß and glucose metabolism, however a regional approach may help us understand the regional impacts of Aß and if, for instance, regional levels of Aß asymmetry drive neural activation asymmetry.

This study supports previously identified relationships between brain asymmetry and neurodegenerative disease pathology. We found that greater left hemisphere asymmetry in the middle frontal gyrus was associated with greater cerebral glucose metabolism. In older cognitively normal adults, we did not identify an association between functional asymmetry and AD risk factors. However, our identification of associations between functional asymmetry and both cognitive function and glucose metabolism indicate that asymmetry may be associated with some early impairment, primarily glucose metabolism which may be an early indicator of preclinical disease.

## Data Availability

Data is available upon request.

## Acknowledgement

This study was supported by P50 AG005133, P01 AG025204, and R37 AG025516 from the National Institute of Health. We would also like to acknowledge and thank the GPN Lab for providing the research infrastructure and continuous mentorship that made this study possible.

